# Generalizability of Tau and Amyloid Plasma Biomarkers in Alzheimer’s Disease Cohorts of Diverse Genetic Ancestries

**DOI:** 10.1101/2024.04.10.24305617

**Authors:** Anthony J. Griswold, Farid Rajabli, Tianjie Gu, Jamie Arvizu, Charles G. Golightly, Patrice L. Whitehead, Kara L. Hamilton-Nelson, Larry D. Adams, Jose Javier Sanchez, Pedro R. Mena, Takiyah D. Starks, Maryenela Illanes-Manrique, Concepcion Silva, William S. Bush, Michael L. Cuccaro, Jeffery M. Vance, Mario R Cornejo-Olivas, Briseida E. Feliciano-Astacio, Goldie S. Byrd, Gary W. Beecham, Jonathan L. Haines, Margaret A. Pericak-Vance

## Abstract

**Introduction:** Plasma phosphorylated threonine-181 of Tau and amyloid beta are biomarkers for differential diagnosis and preclinical detection of Alzheimer disease (AD). Given differences in AD risk across diverse populations, generalizability of existing biomarker data is not assured.

**Methods:** In 2,086 individuals of diverse genetic ancestries (African American, Caribbean Hispanic, and Peruvians) we measured plasma pTau-181 and Aβ42/Aβ40. Differences in biomarkers between cohorts and clinical diagnosis groups and the potential discriminative performance of the two biomarkers were assessed.

**Results:** pTau-181 and Aβ42/Aβ40 were consistent across cohorts. Higher levels of pTau181 were associated with AD while Aβ42/Aβ40 had minimal differences. Correspondingly, pTau-181 had greater predictive value than Aβ42/Aβ40, however, the area under the curve differed between cohorts.

**Discussion:** pTau-181 as a plasma biomarker for clinical AD is generalizable across genetic ancestries, but predictive value may differ. Combining genomic and biomarker data from diverse individuals will increase understanding of genetic risk and refine clinical diagnoses.

## Background

Alzheimer disease (AD) is a progressive and devastating neurodegenerative condition with a prevalence of 5% in individuals 65-74 years of age, increasing to 33% by 85 years of age. This corresponds to 6.7 million people in the US with a current diagnosis of AD, which is estimated to increase to 14 million by 2060.[1] AD has a global impact, with diverse populations around the world affected, but ancestrally diverse groups such as African American (AA) and Hispanic (HI) populations are underrepresented in AD genomic and translational studies[2-11] which in part contributes to health disparities in the US. Compared to Non-Hispanic White (NHW) populations AA populations have higher frequencies of risk factors for complex diseases such as AD, more limited access to health services, poorer health outcomes, and lower life expectancies[12-17] For example, Caribbean HI are twice as likely as non-Hispanic Whites (NHW) to have late-onset AD[18,19] and the incidence of new AD cases in Caribbean HI families is three times the incidence in NHW families.[20] Similarly, compared with NHW individuals, individuals of African ancestry, even from the same local community, are twice as likely to develop AD due to both genetic and environmental influences.[19]

In part, these disparities can be attributed to access to and affordability of AD diagnostic tools such as magnetic resonance imaging (MRI), positron emission tomography (PET) imaging, and cerebrospinal fluid (CSF) assays that can be incorporated as criteria for pre-clinical AD diagnosis.[21] Therefore, the use of AD plasma biomarkers have begun wide use to assist in clinical diagnosis and refine the preclinical stages of AD and MCI.[22-24] These blood-based biomarkers can be used to estimate the underlying biological changes associated with AD diagnosis and prognosis including amyloid deposition (plasma Aβ42/Aβ40 ratio)[25] and tau hyperphosphorylation and tangles (phosphorylated Tau).[26,27]

Critically, however, individuals of African and HI ancestry are underrepresented in most research studies[7-9] including genomic and translational studies,[11,28,29] and particularly for those utilizing AD plasma biomarkers.[30,31] Thus, diverse ancestral groups may not benefit to the same degree as NHW from the clinical applications of genomic and biomarker studies [32-37]. While recent studies have increased representation of African Americans[30,38] and Hispanics,[39,40] the sample sizes of diverse individuals remain a small proportion of the total study cohorts making conclusive statements on the generalizability of these findings difficult.

To address the issues of lack of representation of diverse ancestries in AD biomarker studies, potential differences in AD biomarkers across populations, and the generalizability of AD biomarkers to discriminate between clinically diagnosed AD and cognitively unimpaired (CU) individuals, herein we measured plasma AD related biomarkers pTau-181 and Aβ42/Aβ40 ratio in a large cohort of individuals representing diverse ancestral backgrounds. In these clinically diagnosed cohorts, we compare biomarker concentration between those diagnosed with AD, mild cognitive impairment (MCI), CU across and within the groups. Furthermore, we explore the ability of these biomarkers to discriminate between AD and CU in these diverse sets of individuals. As such, we can better understand the generalizability and utility of these lower-cost and readily available biomarkers for broad use in AD diagnostics in individuals of diverse genetic ancestry.

## Methods

### Ascertainment of Study Participants

Individuals for this study have been ascertained through multiple studies of AD genetics across several sites focused on individuals from underrepresented populations including self-identified Black Americans of African ancestry and Hispanics (Puerto Ricans, Cuban Americans, and Peruvians). The study sites include: the University of Miami (Miami, FL, USA), Wake Forest University (Winston-Salem, NC, USA), Case Western Reserve University (Cleveland, OH, USA), Universidad Central del Caribe (Bayamon, PR, USA), and Instituto Nacional de Ciencias Neurologicas (Lima, Peru). All participants or their proxy provided written informed consent as part of the study protocols approved by the site-specific Institutional Review Boards.

Ascertainment protocols were consistent across sites and clinical data assessments capture sociodemographic information, medical and family history, dementia staging, AD/dementia symptoms, neuropsychological abilities, functional capabilities, and behavioral impairments. Best-estimate diagnoses were generated by a consensus committee consisting of a board-certified neurologist, a physician specializing in dementia, and clinical neuropsychologists, all with expertise in adjudication of AD/dementia. Using all available clinical information, participants were assigned best-estimate clinical diagnoses (AD, MCI, or CU) based on criteria adapted from NIA-Alzheimer’s Association criteria.[21,41,42]

### Genotyping and Assessment of Genetic Ancestry

2,086 total participants were included in this study and classified based on their self-reported race-ethnicity and country of origin as African American (AA), Caribbean Hispanic (Puerto Rican or Cuban American), or Peruvian (PE). For each participant, whole genome genotyping array data on the Illumina Global Screening Array (Illumina, San Diego, CA, USA) was generated. Genotype markers were quality controlled to include only those with high call rate (over 97%) and minor allele frequency over 0.05 within each population. Genetic relatedness was evaluated using the GENESIS R package.[43] Frist, the KING-Robust kinship coefficient estimator was used to calculate the KING matrix that includes pairwise relatedness and measures of pairwise ancestry divergence.[44] The PC-AiR method used ancestry divergence from KING-Robust estimate to perform PCA, which then was used by PC-Relate to estimate pairwise kinship coefficients. Genetically related individuals with closer than 4^th^-degree relatives were identified and only one sample per related group was retained in the analysis. Subsequently, PC-Air was applied to calculate global ancestry using four reference populations were used including AI, EU, AF, and EA from Human Genome Diversity Project (HGDP) data for the reference populations.[45] Admixture proportions were estimated using a model-based clustering algorithm was performed as implemented in the ADMIXTURE software[46] with K=4 for the reference populations.

### Measurement of Plasma AD Biomarkers

Whole blood tubes were collected from the 2,086 participants at the time of study entry by trained phlebotomists. Plasma was isolated from EDTA tubes via centrifugation, aliquoted into polypropylene tubes, and stored at -80°C until further analysis. For a set of African American participants, coagulation was performed in the tube and thus serum was the only available blood fluid available. Plasma/serum concentrations of pTau-181, Aβ42, and Aβ40 were measured using SIMOA chemistry implemented on the Quanterix HD-X analyzer (Quanterix, Billerica, MA, USA)[47] according to manufacturer’s instructions for the pTau-181 Advantage V2 assay and Neurology 3-Plex A assay. Samples were randomized according to age (at time of blood collection), sex, and diagnosis and assayed in duplicate on each Simoa plate. Initial data analysis was performed using the Quanterix Analyzer v1.6 software to calculate standard curves and biomarker concentrations. After quality control including removing of related participants, failed biomarker assays, and removal of outliers greater than three standard deviations from the mean 1,368 participants with plasma and 721 participants with serum remained in the analyses (Table 1).

**Table 1.** Characteristics of the study participants and biomarker levels.

| Alzheimer's Disease | All samples<br>(N=421) | African<br>American<br>(N=71) | Caribbean<br>Hispanic<br>(N=312) | Peruvian<br>(N=38) |
| --- | --- | --- | --- | --- |
| Sex |  |  |  |  |
| Female | 283 (67.2%) | 47 (66.2%) | 209 (67.0%) | 27 (71.1%) |
| Male | 138 (32.8%) | 24 (33.8%) | 103 (33.0%) | 11 (28.9%) |
| Age (years) |  |  |  |  |
| Mean (SD) | 79.4 (7.85) | 80.0 (8.48) | 79.3 (7.76) | 79.8 (7.59) |
| pTau181 level (pg/mL) |  |  |  |  |
| Mean (SD) | 2.76 (1.81) | 3.00 (1.49) | 2.68 (1.87) | 3.08 (1.81) |
| Aβ42/40 | | | | |
| Mean (SD) | 0.0410 (0.0137) | 0.0478 (0.0159) | 0.0402 (0.0130) | 0.0352 (0.00990) |
| Mildly Cognitive<br>Impaired | All samples<br>(N=270) | African<br>American<br>(N=88) | Caribbean<br>Hispanic<br>(N=182) | Peruvian<br>(N=0) |
| Sex |  |  |  | - |
| Female | 193 (71.5%) | 67 (76.1%) | 126 (69.2%) | - |
| Male | 77 (28.5%) | 21 (23.9%) | 56 (30.8%) | - |
| Age (years) |  |  |  | - |
| Mean (SD) | 74.9 (7.36) | 72.7 (7.36) | 75.9 (7.15) | - |
| pTau181 level (pg/mL) |  |  |  | - |
| Mean (SD) | 1.91 (1.26) | 1.90 (1.46) | 1.92 (1.15) | - |
| Aβ42/40 | | | | - |
| Mean (SD) | 0.0455 (0.0149) | 0.0536 (0.0168) | 0.0418 (0.0123) | - |
| Cognitively<br>Unimpaired | All samples<br>(N=677) | African<br>American<br>(N=234) | Caribbean<br>Hispanic<br>(N=355) | Peruvian<br>(N=88) |
| Sex |  |  |  |  |
| Female | 533 (78.7%) | 197 (84.2%) | 270 (76.1%) | 66 (75.0%) |
| Male | 144 (21.3%) | 37 (15.8%) | 85 (23.9%) | 22 (25.0%) |
| Age (years) |  |  |  |  |
| Mean (SD) | 73.2 (7.23) | 70.9 (6.73) | 74.7 (7.36) | 72.9 (6.41) |
| pTau181 level (pg/mL) |  |  |  |  |
| Mean (SD) | 1.98 (2.60) | 2.02 (4.15) | 1.84 (1.05) | 2.40 (1.22) |
| Aβ42/40 | | | | |
| Mean (SD) | 0.0477 (0.0247) | 0.0575 (0.0196) | 0.0432 (0.0275) | 0.0408 (0.0138) |

African American Serum Samples
|  | CU<br>(N=370) | MCI<br>(N=103) | AD<br>(N=248) | Overall<br>(N=721) |
| --- | --- | --- | --- | --- |
| Sex |  |  |  |  |
| Female | 275 (74.3%) | 81 (78.6%) | 194 (78.2%) | 550 (76.3%) |
| Male | 95 (25.7%) | 22 (21.4%) | 54 (21.8%) | 171 (23.7%) |
| Age (years) |  |  |  |  |
| Mean (SD) | 69.5 (6.74) | 73.6 (7.03) | 80.8 (8.12) | 74.0 (8.89) |
| pTau181 level (pg/mL) |  |  |  |  |
| Mean (SD) | 1.50 (1.46) | 1.18 (1.22) | 2.69 (2.26) | 1.86 (1.85) |
| Aβ42/40 | | | | |
| Mean (SD) | 0.0542 (0.113) | 0.0481 (0.0182) | 0.0491 (0.0458) | 0.0519 (0.0899) |

### Biomarker Statistical Analysis

Biomarker concentrations were log10-transformed to satisfy normality assumptions in the downstream analysis. The differences in biomarker levels across ancestral groups and between AD, MCI and CU individuals was analyzed using a linear regression model adjusted for age, sex, and population substructure (principal components 1-3) followed by the post hoc least significant difference test for pairwise group comparison and adjusted for multiple comparisons by Bonferroni.

To assess the diagnostic performance and construct receiver operator characteristic (ROC) curves, we employed three logistic regression models: pTau-181 concentration alone, Aβ42/Aβ40 alone, and pTau-181 concentration and Aβ42/Aβ40 together. For this analysis we used only individuals having data available on both pTau-181 and Aβ42/Aβ40 biomarkers (N = 1,298 for plasma, N = 260 for serum). Area under the curves (AUC) of the three models were compared using the DeLong test and adjusted for multiple comparisons (Bonferroni). All analysis were performed using R v.4.1.1 with packages: lmerTest,[48] multcomp,[49] lsmeans,[50] and pROC,[51] and visualizations created with ggpubr.[52]

## Results

### Global Genetic Ancestry of Participants

As detailed in Table 1, the participants in this study were ascertained as part of four distinct cohorts: African American (AA), and HI with origins from the Caribbean (CH - Puerto Rican or Cuban American) or Peru (PE). Figure 1A illustrates principal component analysis results of the study samples and four continental populations from the HGDP reference panel. Genomic admixture varied widely across cohorts with respect to proportions of European, African, Amerindian, and East Asian ancestries (Figure 1B). Given similarity of ancestral admixture between Puerto Ricans and Cuban Americans and the relatively small sample size of Cuban Americans, these cohorts were combined in further analysis as Caribbean Hispanics. Global ancestry analysis confirmed primarily two-way admixture in AA (African and European), three-way admixture in CH (African, European, and Amerindian), and four-way admixture in PE (African, European, Amerindian, and East Asian) with a high proportion of Amerindian background.

**Figure 1.**
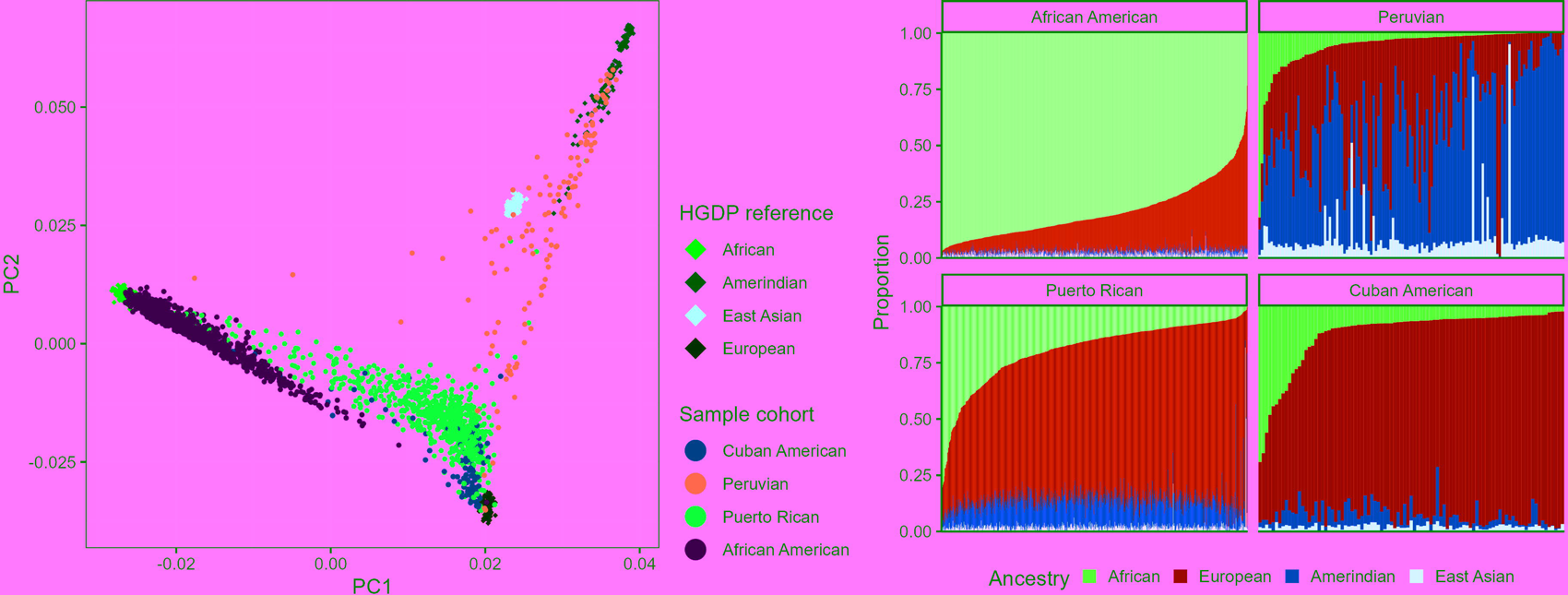
Global genomic ancestry analysis. **A)** Principal component analyses. Estimation of relationship to ancestral reference groups from Human Genome Diversity Project (HGDP): Maroon = African, Green = Amerindian, Yellow = East Asian, Blue = European, or each sample cohort: Light Green = Cuban American, Violet = Peruvian, Red = Puerto Rican, Aqua = African American. **B)** Ancestral admixture estimations. Each column on the X-axis represents one participant. Colors in each vertical line represent the proportion of ancestral admixture. Red = African, Blue = European, Green = Amerindian, Yellow = East Asian.

### Plasma pTau-181 and Aβ42/Aβ40 ratios across ancestries

We first set out to determine how AD plasma biomarker levels of pTau-181 and Aβ42/Aβ40 compare across ancestries, within each diagnostic category of AD or CU. In general, plasma pTau-181 concentrations were consistent across these populations. There was a modest but significant increase in Peruvian controls relative to the other two groups (p=0.02 vs. AA, p=0.002 vs. CH), Figure 2A), but among AD there were no significant differences between groups (Figure 2B). The Aβ42/Aβ40 ratio was significantly higher in African Americans relative to the other two groups in the CU comparison p=2.38x10^-7^ vs Peurivan, p=4.36x10^-7^ vs CHI, Figure 2C) and also trending higher in AD with a significant difference between AA and CH (p = 0.05, Figure 2D).

**Figure 2.**
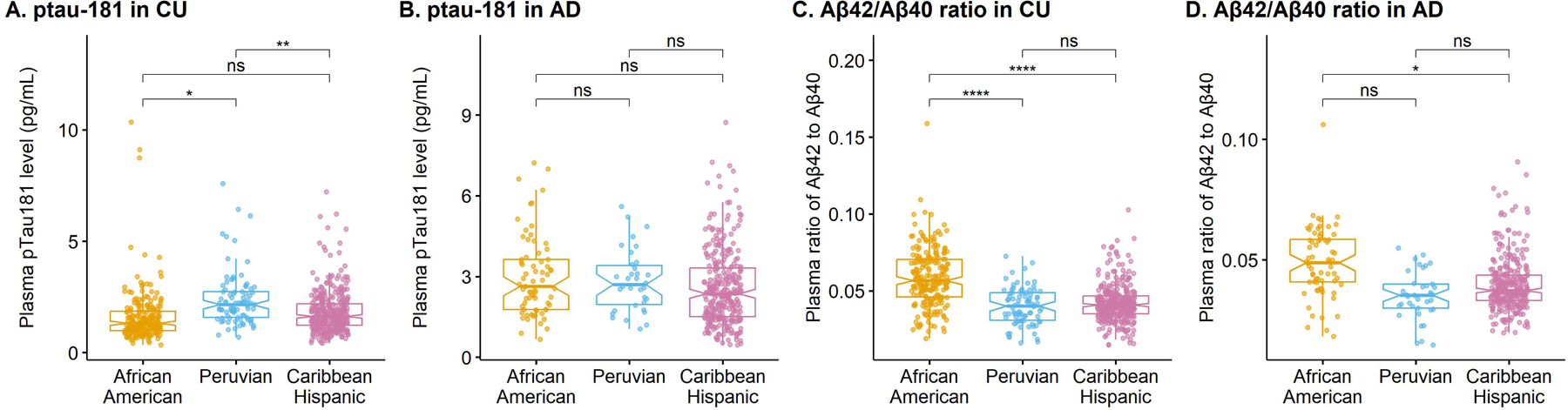
AD biomarker levels across ancestral groups African Americans, Peruvians, and Caribbean Hispanics. **A)** pTau-181 in cognitively unimpaired. **B)** pTau-181 in AD. **C)** Aβ42/Aβ40 in cognitively unimpaired. **D)** Aβ42/Aβ40 in AD. **** p<0.0001, ***p<0.001, **p<0.01, *p<0.05, ns = not significant

### Plasma pTau-181 and Aβ42/Aβ40 comparisons

Plasma pTau-181 concentrations were significantly increased in individuals with diagnosed clinical AD compared to CU and MCI (p = 1.67x10^-13^, Figure 3A) considering all individuals across all cohorts (Figure 3A). When analyzing each cohort separately, plasma pTau-181 concentrations were significantly higher in AD vs CU in AA (p = 1.88x10^-8^, Figure 3B) and CH (p = 3.13x10^-7^, Figure 3D) with a trend that did not reach statistical significance for Peruvians (p = 0.07, Figure 3C). When considering AD vs MCI, there was again a statistically higher pTau-181 concentration in overall (p = 7.83x10^-8^, Figure 3A) as well AA (p = 7.95x10^-5^, Figure 3B) and CH (p = 5.25x10^-5^, Figure 3D). The plasma Aβ42/Aβ40 ratio was significantly lower in AD of AA relative to CU (p = 0.02, Figure 4A-D). There was no statistically significant difference in the concentration of pTau-181 nor Aβ42/Aβ40 ratio between MCI and CU when considering all data together or when looking at specific cohorts individually.

**Figure 3.**
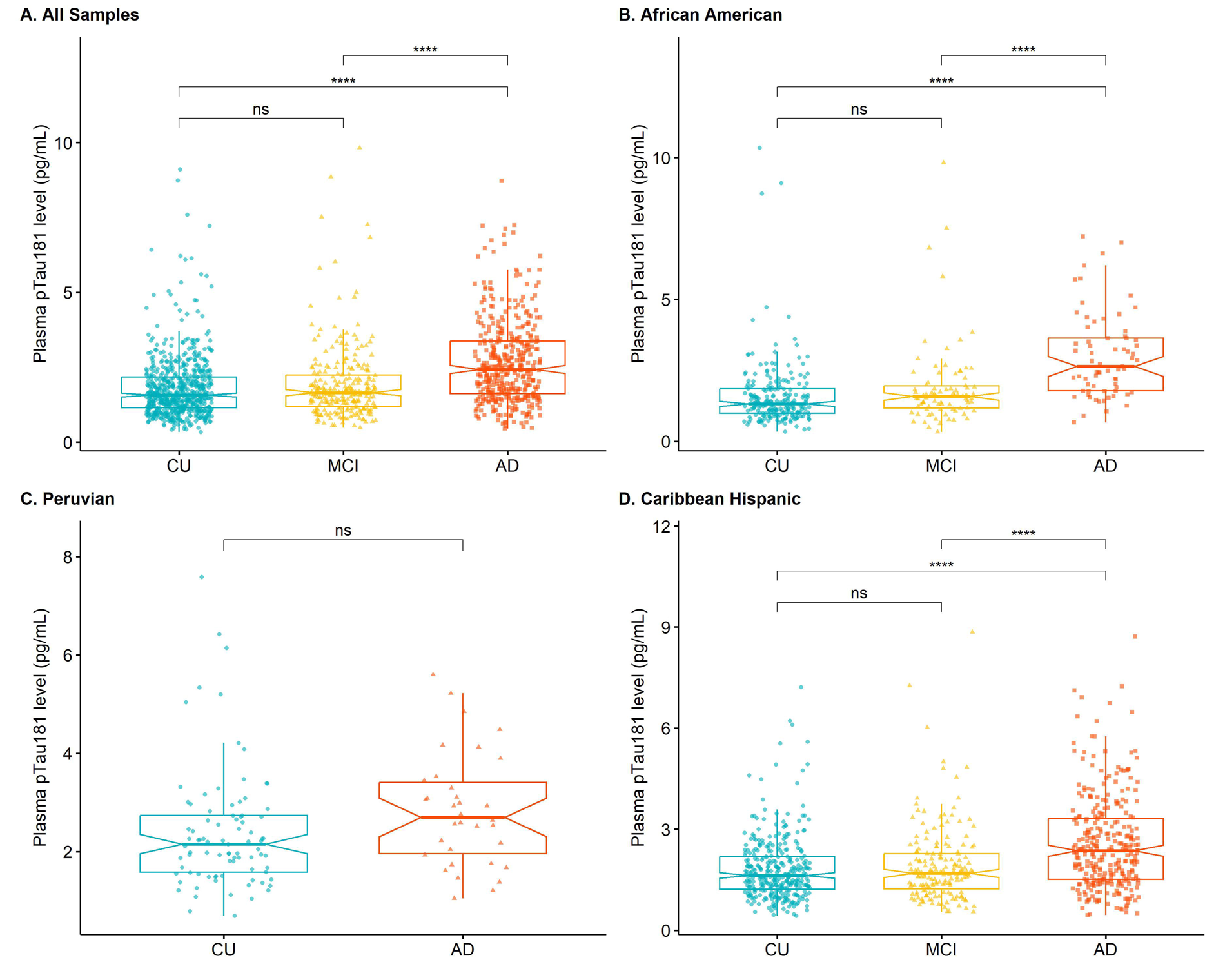
Plasma pTau-181 comparison analysis. Comparisons of the pTau-181 concentration in clinical diagnosis statuses of Alzheimer Disease (AD), mild cognitively impaired (MCI), or cognitively unimpaired (CU) in: **A)** The overall combined cohort, **B)** African Americans, **C)** Peruvians, and **D)** Caribbean Hispanics. Each point represents one individual’s pTau-181 measurement. Concentrations are shown as median ± interquartile range, **** p<0.0001, ***p<0.001, **p<0.01, *p<0.05, ns = not significant

**Figure 4.**
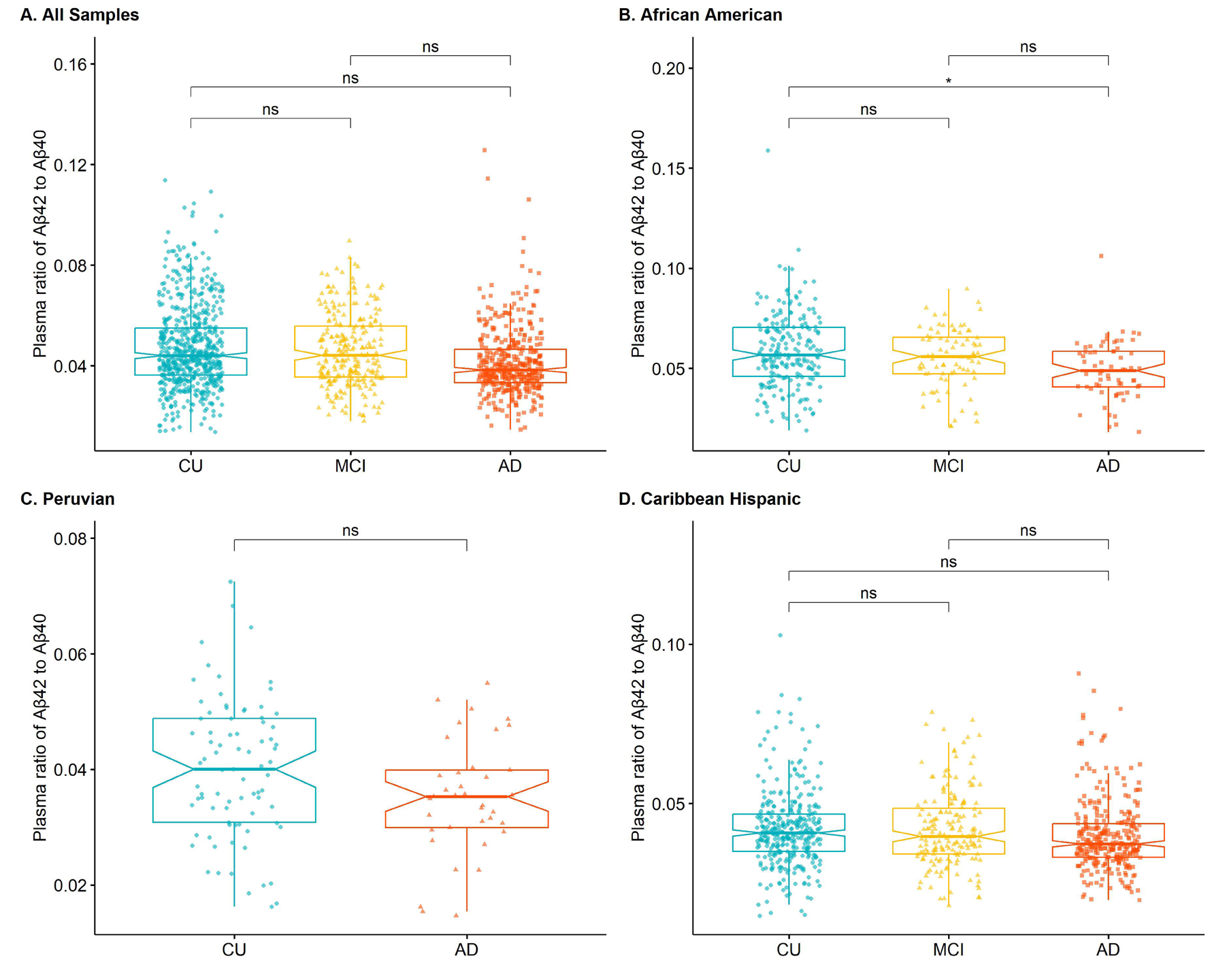
Plasma Aβ42/Aβ40 ratio comparison analysis. Comparisons of the Aβ42/Aβ40 ratio in clinical diagnosis statuses of Alzheimer Disease (AD), mild cognitively impaired (MCI), or cognitively unimpaired (CU) in: **A)** The overall combined cohort, **B)** African Americans, **C)** Peruvians, and **D)** Caribbean Hispanics. Each point represents one individual’s Aβ42/Aβ40 ratio. Ratios are shown as median ± interquartile range, *p<0.05, ns = not significant

### Serum pTau-181 and Aβ42/Aβ40 comparisons

We analyzed the biomarkers in a subset of African ancestry individuals for whom only serum, rather than plasma, was available. We first evaluated potential differences between serum and plasma pTau-181 and Aβ42/Aβ40 ratio within each diagnostic category for African Americans. Plasma pTau-181 concentration was higher than serum in both CU (p = 9.16x10^-6^, Figure 5A) and AD (p = 1.63x10^-3^, Figure 5B). The Aβ42/Aβ40 ratio was lower in serum in both CU (p = 2.44x10^-13^, Figure 5C) and AD (p = 0.05, Figure 5D). Given these differences, we performed analysis within these serum samples separately. Like the plasma analysis, serum pTau-181 concentrations were significantly increased in individuals with diagnosed clinical AD compared to either CU or MCI (Figure 6A). Also, the serum Aβ42/Aβ40 ratio did not significantly differ (Figure 6B).

**Figure 5.**
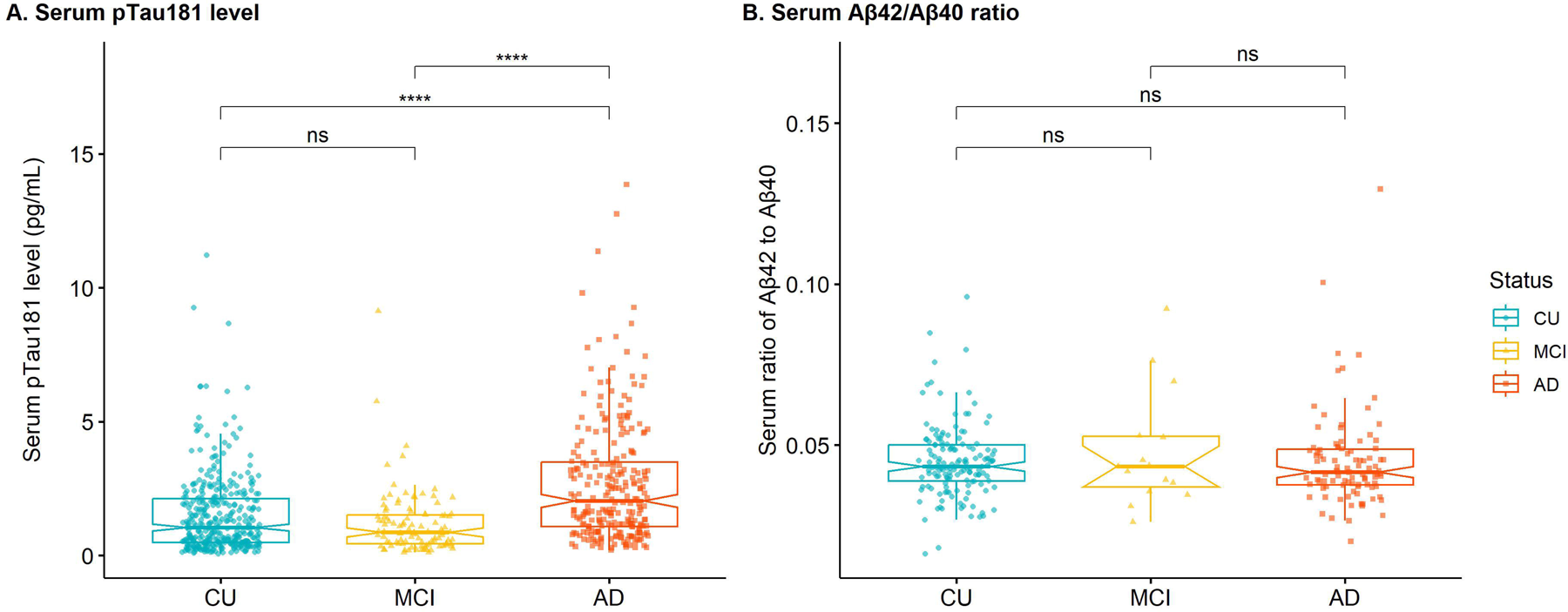
pTau-181 and Aβ42/Aβ40 ratio comparisons in African American plasma and serum analysis. Comparisons of pTau-181 concentrations between serum and plasma in **A)** cognitively unimpaired (CU) and **B)** Alzheimer Disease (AD). Aβ42/Aβ40 ratio between serum and plasma in **C)** cognitively unimpaired (CU) and **D)** Alzheimer Disease (AD). Each point represents one individual’s measurement. Concentrations are shown as median ± interquartile range. **** p<0.0001, **p<0.01, *p<0.05

**Figure 6.**
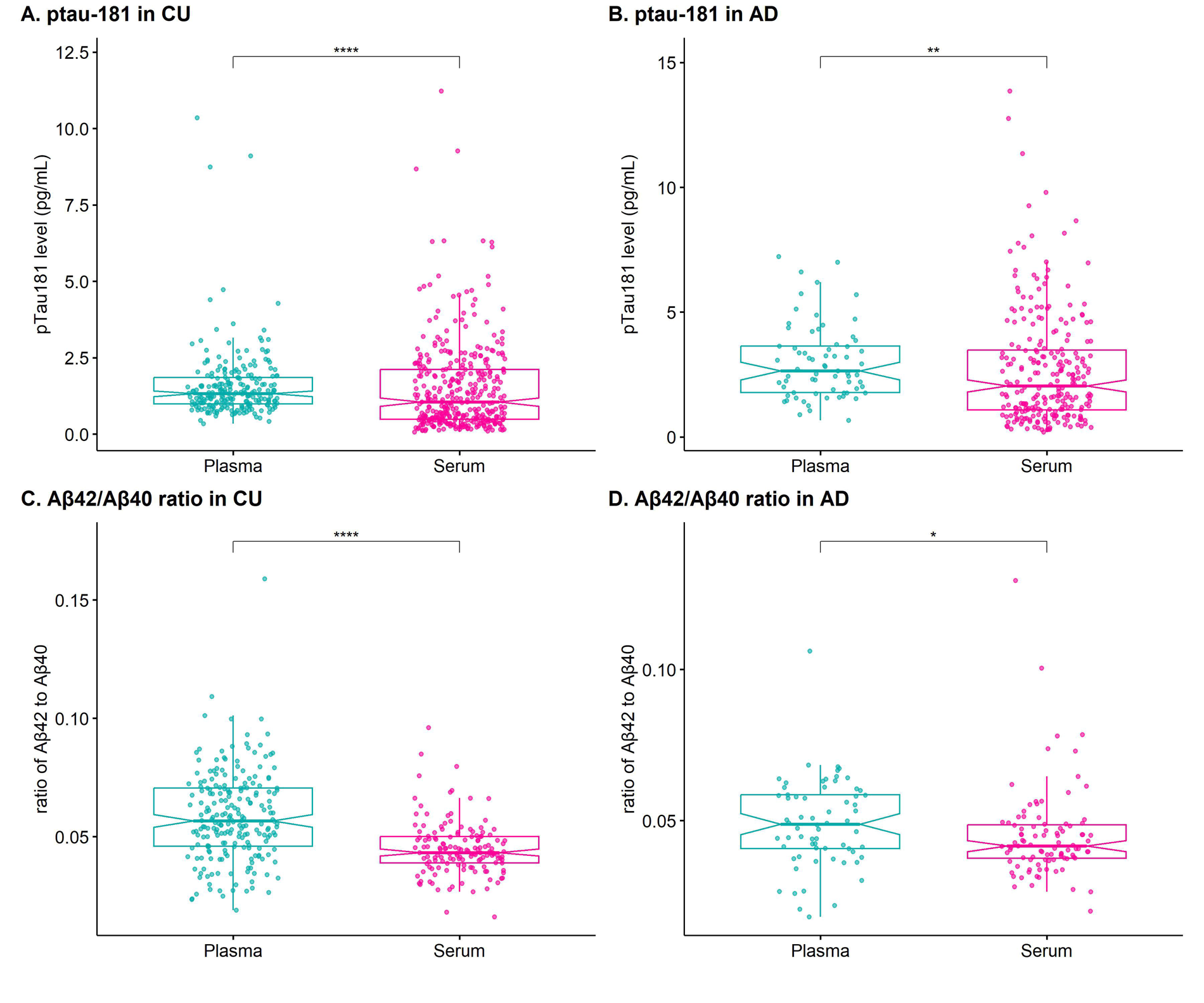
Serum pTau-181 and Aβ42/Aβ40 ratio comparison analysis. Comparisons of the **A)** pTau 181 concentration and **B)** Aβ42/Aβ40 ratio in clinical diagnosis statuses of Alzheimer Disease (AD), mild cognitively impaired (MCI), or cognitively unimpaired (CU) in a subset of African American individuals for which plasma was not available. Each point represents one individual’s measurement. Concentrations are shown as median ± interquartile range. ****p < 0.0001, ns – not significant.

### Discriminatory analysis of plasma and serum pTau-181 and Aβ42/Aβ40

We used logistic regression models to assess the accuracy of plasma and serum pTau-181 concentration and Aβ42/Aβ40 ratio in predicting clinical disease status. We evaluated the performance of each biomarker individually and then in combination using the area under the receiver operating characteristic curve (AROC) for each model (Figure 7). In general, pTau-181 was better at predicting status than Aβ42/Aβ40 ratio, and the classification improved slightly when both biomarkers were used together. Notably, the accuracy varies over the individual cohorts with combined AROC ranging from 0.85 in plasma measurements for African Americans to 0.65 in Peruvians.

**Figure 7.**
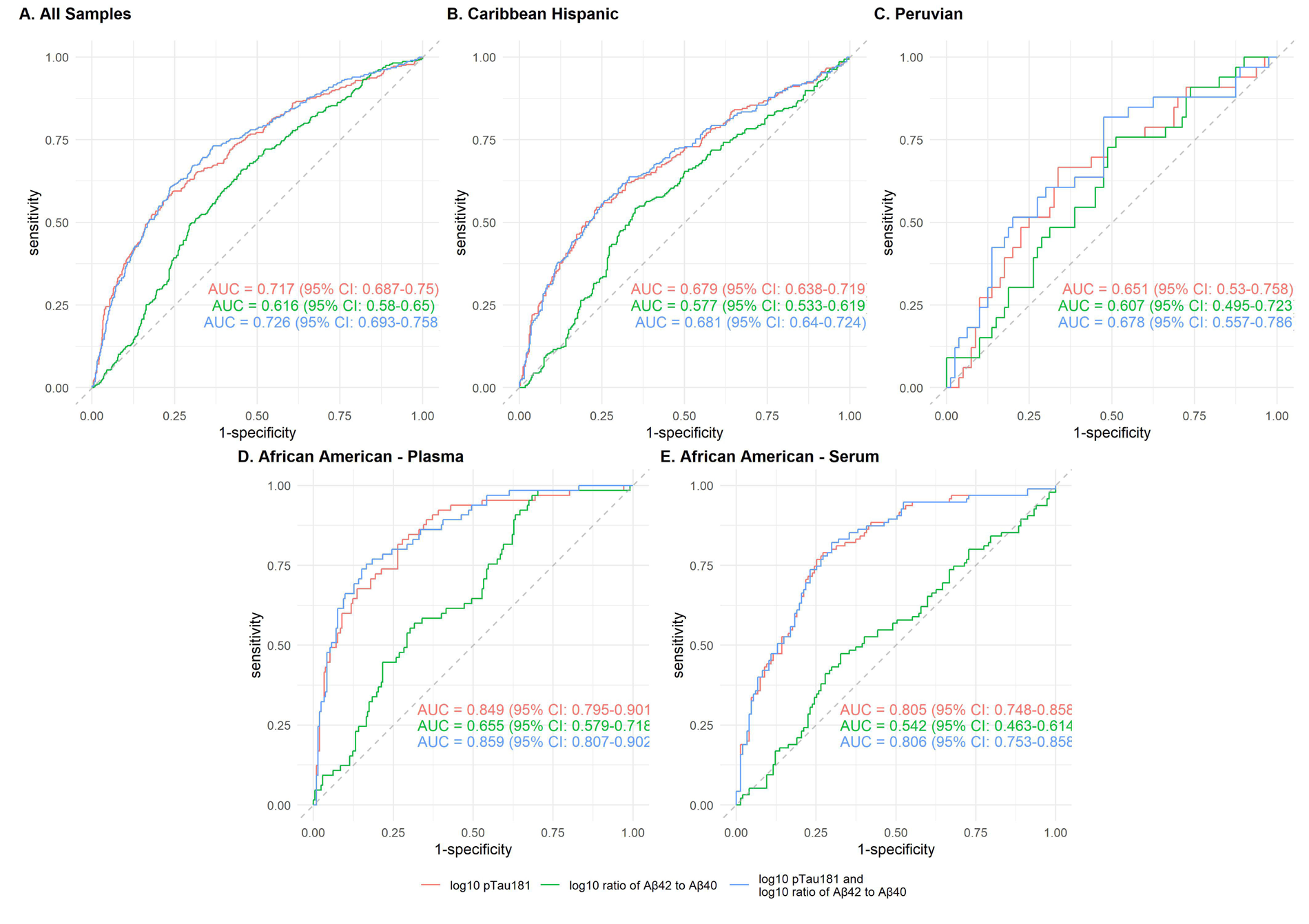
Receiver operating characteristic (ROC) curves of plasma and serum biomarkers for the classification of Alzheimer disease status in: **A)** The overall combined cohort, **B)** Caribbean Hispanics, **C)** Peruvians, **D)** African Americans - plasma, **E)** African Americans - serum. Red lines represent pTau-181 alone, green lines Aβ42/Aβ40 ratio alone, and blue lines the combination of pTau-181 and Aβ42/Aβ40 ratio.

## Discussion

Herein, we have performed the largest to date study of plasma biomarkers in diverse ancestral AD cohorts. Importantly, we found that the plasma concentrations of pTau-181 are generally consistent across populations within clinical diagnostic categories, with a small elevation in Peruvian CU relative to African Americans and Caribbean Hispanics, while the Aβ42/Aβ40 ratio was higher in African American AD and CU cohorts relative to Peruvians or Caribbean Hispanics. Moreover, we found that the plasma/serum concentration of pTau-181 was significantly greater in AD relative to CU and MCI across these diverse ancestral cohorts. We did not detect differences between the statuses when comparing the Aβ42/Aβ40 ratio across or within the cohorts. These correspond to pTau-181 offering strong predictive value of AD status in these data, with Aβ42/Aβ40 adding little more to the model. Interestingly, however, the predictive values varied widely across the ancestral groups ranging from 0.85 (AA) to 0.65 (PE).

The pTau-181 results generally reflect work done in previous large cohort studies done primarily in European non-Hispanic White (NHW) individuals showing it to be strongly associated with AD status[27]. Here we demonstrate generalizability of the pTau-181 biomarker in large cohorts of clinically diagnosed individuals without stratification by alternative biomarkers such as PET imaging as had previously been described for NHW.[53] Conversely, the Aβ42/Aβ40 ratio did not offer strong association with AD status despite several previous reports of this association. While we cannot dismiss the possibility that this is due to either ancestral diversity or the clinical diagnosis used in this study, it has been previously shown that the SIMOA chemistry assays for these analytes may lack the sensitivity required to detect such differences [54]. In biomarker studies of more diverse individuals, the utility of pTau-181 has been shown in predicting AD status in Caribbean Hispanics[39] as well as African Americans.[38,55] However, the larger sample sizes of diverse cohorts reported here, 3-5 times larger than previous, indicate that this is a robust and reproducible observation; that pTau-181 can be incorporated in more diverse populations as a potential aide in diagnostic prediction. However, it is clear that this biomarker lacks complete discrimination of status as there is significant overlap in individual values between AD, MCI, and CU. This suggests that pTau-181 should be combined with other clinical tests and existing risk factors in making diagnostic decisions or could be used as longitudinal measure of rate of change in progression toward dementia.

While pTau-181 was significantly higher in AD than MCI or CU across the diverse cohorts investigated, there was a range of discriminatory AOC values represented. Notably in the Peruvian cohort, the overall AROC value was lower (0.64) than in the other cohorts. The underlying reason for this cannot be determined, though given the higher overall pTau-181 levels in Peruvian CU is likely to affect this analysis. Intriguingly, this may be related to admixture of genetic ancestries altering the utility of biomarkers in these cohorts. Further investigation related to the complex interplay between genetics, social and environmental determinants of health, and AD biomarkers will be critical to continue to assess these questions.

Overall, we provide further evidence that AD plasma biomarkers, particularly pTau-181, can be used to discriminate between AD and MCI or CU not only in NHW populations but in ancestrally diverse cohorts. We have added significantly to the ongoing research in this area while greatly increasing the inclusivity of individuals that have traditionally made up only a very small proportion of participants in biomarker studies. Further confirmation of these observations in diverse cohorts including either autopsy confirmed individuals or those with extensive diagnostic imaging could provide further validation of the use of these biomarkers in clinical based cohorts while assisting in establishing ancestry specific cut-points for diagnostic purposes. Ultimately, adding in AD plasma biomarkers with genetic and environmental information in these diverse populations will continue to reveal the general and ancestry specific risk profile of AD aiding in development of precision treatment strategies.

## Data Availability

All data produced in the present study are available upon reasonable request to the authors.

## Consent Statement

All participants, or their proxy, provided written informed consent as part of the study protocols approved by the site-specific Institutional Review Boards.

## Acknowledgements

We acknowledge the participation of the individuals included in this study. Biomarker data was generated in collaboration with the Center for Genome Technology (CGT) from the John P. Hussman Institute for Human Genomics (HIHG).

## Funding

This work was supported by the National Institutes of Health National Institute on Aging (grant numbers AG070935 to A.J.G and AG072547, AG052410, AG070864, and AG074865 to M.A.P.V.)

## Conflict of Interest

The authors declare no conflicts of interest.

## Notes

### Competing Interest Statement

The authors have declared no competing interest.

### Author Declarations

All participants, or their proxy, provided written informed consent as part of the study protocols approved by the site-specific Institutional Review Boards at the University of Miami, Case Western Reserve University, Wake Forest University, Instituto Nacional de Ciencias Neurologicas (Lima, Peru), or Universidad Central Del Caribe (Bayamon, Puerto Rico)

